# Microsampling for monitoring gentamicin in neonates

**DOI:** 10.1101/2021.03.27.21254449

**Authors:** Suzanne L. Parker, Adam D. Irwin, Francine Hosking, Deanne August, Brittany Schoenmaker, Saurabh Pandey, Steven C. Wallis, Jeffrey Lipman, Jason A. Roberts, Mark W Davies

## Abstract

Gentamicin is recommended as first-line treatment of neonatal sepsis. The use of gentamicin is associated with toxicity which complicates neonatal dosing and necessitates therapeutic drug monitoring (TDM).

In a proof-of-concept investigation, we sought to compare (1) gentamicin concentrations obtained using volumetric absorptive microsampling (VAMS) to standard TDM plasma samples, and (2) the time taken to report results obtained using VAMS compared to standard TDM by the local hospital chemical pathology service.

The difference between gentamicin concentrations obtained from plasma collected for routine clinical care and calculated plasma concentrations, based on samples collected in whole blood using VAMS, was −18.0% and −0.4% for two patients. The research laboratory reported results within the time taken for the routine chemical pathology laboratory to report results. This proof-of-concept study demonstrates that the use of microsampling for TDM by pathology services can fulfil the requirements of providing an accurate gentamicin concentration in a timely manner.

## Introduction

Neonatal sepsis is a significant contributor to global mortality, with recent studies suggesting an increasing incidence, driven by increasing intervention in pre-term neonates[1]. Gram-negative organisms such as *Escherichia coli* and *Klebsiella pneumoniae* increasingly predominate and are frequently associated with antimicrobial resistance[2]. It is imperative that recommended and currently available antibiotics are used optimally in this most vulnerable group.

Gentamicin is recommended by the World Health Organisation as a first-line antimicrobial for neonatal sepsis, and is the most commonly prescribed antimicrobial in neonatal sepsis[3]. It is used to treat invasive Gram-negative infections in neonates and associated with nephrotoxicity and ototoxicity[4]. Gentamicin is a prototypical therapeutic drug monitoring (TDM) agent with trough concentrations below 2 mg/L a common target. Blood sampling in very small and premature neonates presents a complex challenge where, relative to body size, the large volumes of blood that are collected for routine clinical care can cause anaemia requiring blood transfusions.

Bioanalytical methodology used for measuring gentamicin concentrations in TDM should be reliable, accurate and able to provide a result in a timely manner. Recent advances in bioanalysis have led to interest in reducing the volume of blood sampled from the patient to less than 0.05 mL (microsampling) to support clinical care. The volumetric absorptive microsampling (VAMS) device, marketed as Mitra® by Neoteryx, collects a known volume (10.8 µL) of whole blood onto a hydrophilic tip. This sample can be used in quantitative analysis either while the blood remains wet or the tip can be dried and stored.

This proof-of-concept investigation sought to compare (1) the gentamicin concentrations obtained using VAMS against the results obtained from the TDM plasma sample, and (2) the time taken for analysis of the VAMS compared to routine TDM by the local hospital chemical pathology service.

## Methods

A prospective proof-of-concept study involving two neonates was conducted within a clinical correlative study (the OPTIONS study) at the neonatal intensive care unit of a tertiary hospital in 2020. A standard protocol for dosing neonates in the unit is the administration of gentamicin as a 3.5 mg/kg dose every 36 h for patients less than 30 weeks gestational age (GA) and every 24 h for patients greater than or equal to 30 weeks gestational age. Dose adjustments are made based on TDM performed at least 30 minutes prior to the third dose. If the trough concentration is ≥ 2 mg/L and gentamicin treatment is to continue, the dosing interval is then prolonged, with the TDM repeated prior to the administration of the next gentamicin dose. Patient information for the study was collected from medical records and chemical pathology laboratory and pharmacy databases. The study was approved by the Royal Brisbane & Women’s Hospital human research ethics committee (HREC/2020/QRBW/55401). Informed consent was obtained from parents/guardians for participants in this study and for publication of the results.

The acceptance criteria applied to the comparison of calculated plasma concentrations (Cp*) derived from VAMS to the measured plasma (Cp) results was that the difference between the two results must be within 20% of the mean of the measured (Cp) and calculated plasma concentrations (Cp*). A brief description of the blood collection, validation results and analytical technique is presented in the Supplementary Material. The nurses assigned to the care of each of the neonatal patients collected the VAMS; both nurses having received prior training on the correct collection process.

## Results

Two neonatal patients were recruited into this study and provided both TDM samples, as part of routine clinical care, and whole blood samples using a VAMS device. One patient was extremely pre-term, with a weight of 664 g and serum creatinine of 82 µmol/L. The other patient was full term, with a weight 3330 g and serum creatinine of 88 µmol/L. Both patients were receiving gentamicin for presumptive treatment for sepsis. When patients were recruited into the study the research bioanalytical laboratory was notified of the time that samples were to be collected, providing time for the laboratory to prepare for the analysis. The results of the measurement of gentamicin concentrations are reported in Table 1. Quality assurance data from the analytical batches is presented in the Supplemental Material.

**Table 1:** Neonatal patient gentamicin concentrations obtained from paired plasma TDM samples and VAMS, with the calculated plasma concentration (Cp*) being the gentamicin result reported to the clinic when using volumetric absorptive microsampling (VAMS)

| Patient weight (g) | Plasma sample (Cp) |  | VAMS (Cv) |  | Haematocrit (%) | Calculated plasma concentration from Cv (Cp*)<sup>a</sup> | % difference between Cp and Cp* |
| --- | --- | --- | --- | --- | --- | --- | --- |
|  | Mean (mg/L, n = 3) | Imprecision (%) | Mean (mg/L, n = 2) | Imprecision (%) |  | (mg/L) |  |
| 664 | 0.896 | 6.1 | 0.421 | 12.9 | 0.438 | 0.749 | -18.0 |
| 3330 | 2.76 | 5.5 | 1.17 | 14.5 | 0.573 | 2.74 | -0.4 |

<sup>a</sup> Calculation for Cp\* = Cv / (1 – Hct)
GA: gestational age;
Cp: measured plasma concentration;
Cv: measured concentration in whole blood on VAMS;
Hct: haematocrit;
Cp\*: calculated plasma concentration (using equation a)

Each patient provided TDM samples and VAMS approximately one hour prior to their scheduled gentamicin dose. The time to report the gentamicin concentrations was 2 h 55 min and 2 h 33 min for the pathology laboratory, and 1 h 0 min and 2 h 8 min for the research bioanalytical laboratory for the first and second patient, respectively.

## Discussion

In our study, the results of gentamicin plasma concentrations obtained using VAMS correlated with those collected from the TDM plasma sample. Furthermore, for each for the two patients, the analysis performed by the research bioanalytical laboratory was able to provide a result as quickly as that obtained from the pathology laboratory.

This is the first paper to describe the clinical application of microsampling to measure gentamicin concentrations. There is one published study using VAMS for patient monitoring, where the authors reported finding an excellent agreement between VAMS and standard blood samples collected for measuring HbA1c[5].

The results of this study comply with the requirements of an incurred sample reanalysis test as described in bioanalytical validation guidelines provided by the U.S. FDA. This test is usually applied to the repeat analysis of a single sample in bioanalysis, whereas our analysis is performed on two sample types (whole blood and plasma). Meeting this test criteria supports our conclusion that there is a reasonable correlation between the calculated plasma concentrations derived from VAMS and the measured plasma concentrations. Further research is required to complete full correlative or ‘bridging’ studies to adhere to the regulatory requirements for the use of bioanalytical technologies.

An important feature of TDM is the ability of a method to provide a result to a clinical unit in a timely manner. Based on the results of this study it is feasible that this sampling technique and methodology could be applied to routine TDM of gentamicin.

Gentamicin has been found to partition slowly intracellularly and at low concentrations. On the basis that most neonates receive only a few days of gentamicin treatment, our equation to convert concentrations from whole blood to plasma (see Supplemental Material) applies the patient haematocrit only and does not include partitioning into the cellular component. The translation of VAMS for TDM would require a simple change in practice by a pathology laboratory with the use of the conversion equation and the continued reporting of plasma concentrations. This change would not impact on clinical targets or the interpretation of the gentamicin concentration results in the clinic.

This, to the best of our knowledge, is the first report of the application of VAMS to the TDM of gentamicin in neonates. This proof-of-concept study demonstrates the translation of VAMS for TDM of gentamicin for clinical care can fulfil the requirements of providing an accurate and reliable concentration in a timely manner. The benefit to the patient of low-volume blood microsampling for reducing the burden of routine clinical care of neonates is high. Further research is required to support the application of microsampling to improve patient care for TDM of gentamicin.

## Data Availability

All data relevant to the study are included in the article or uploaded as supplementary information

## Transparency Declarations

None to declare.

