## Supplemental File for "Microsampling for monitoring gentamicin in neonates"

**Supplemental Material:**

**Sample collection and analysis:**

Blood samples were collected from patients for TDM using a 1.0 mL syringe from an indwelling catheter or by heel-prick. Blood samples were collected onto the VAMS either from the same syringe used to collect the TDM sample, by delivery of blood onto a small plastic tray, or from the same heel-prick used to collect the TDM sample. The VAMS were collected and taken to a local research bioanalytical laboratory for immediate analysis. After TDM had been performed by the pathology laboratory the patient plasma samples were retrieved and stored for subsequent analysis. The VAMS were assayed in duplicate immediately on collection at the research bioanalytical laboratory. The plasma TDM sample was assayed in triplicate across two days, with one analysis performed on the first day and a duplicate analysis on the second day to provide an inter-day precision analysis of the analytical methodology. The analysis in the research bioanalytical laboratory was performed in batches that contained calibration and quality control samples, the clinical sample and a drug-free matrix-matched sample, in the same manner as batch analysis is routinely performed in a chemical pathology laboratory.

**Data analysis:**

The conversion of gentamicin concentrations from whole blood to plasma was performed using the following equation:

Cp* = Cv / (1 - Hct)

Where Cp* is the calculated plasma concentration, Cv is the measured concentration in whole blood from VAMS, and Hct is the patient’s haematocrit. This equation is appropriate when the drug is sequestered into the plasma portion of the blood, as is the case for gentamicin. The difference between the TDM plasma sample and VAMS concentrations were calculated using Microsoft® Excel for Mac version 15.23.1.

**Validation data summary of VAMS:**

Calibration standards were prepared in whole blood and applied to VAMS. Inter-assay analysis of a linear calibration without weighting (n = 5) produced a slope of (mean ± SD) 0.135 ± 0.028 and intercept of 0.002 ± 0.024. On three occasions prior to the study the calibration was performed across the range of 0.2 to 20 mg/L, mean correlation coefficient of 0.997, and on the study days a truncated calibration was performed of 0.6 to 2 mg/L.

Quality control samples prepared at 0.6 and 2.0 mg/L in whole blood and applied to VAMS. An inter-assay analysis provided an accuracy of 108 % and 101 % and imprecision of 11.2 % and 5.1% at 0.6 and 2.0 mg/L, respectively.

**Validation data summary of plasma samples:**

Calibration standards were prepared in plasma across a concentration range of 0.2 to 20 mg/L. A quadratic calibration (n = 3 occasions) provided the best fit of the data, using (1/x^2^) weighting produced a slope of 1.76 ± 1.97, intercept -0.039 ± 0.066, quadratic term 0.014 ± 0.011 and correlation coefficient of 0.996 ± 0.002.

Quality control samples were prepared at 0.6, 2.0 and 16 mg/L in plasma with inter-assay precision calculated at 8.6, 9.3 and 5.8 % and accuracy 108, 107 and 99.3 %. Of the 21 quality control samples assayed in the analytical batches, 19 met the acceptance criteria of back-calculated concentrations being within ± 15%. All batches met batch acceptance criteria with no more than two and not more than one at each concentration of the quality control samples failed to meet batch acceptance criteria.

**Sample preparation:**

On collection, the wet VAMS was immediately transferred into a 1.8 mL microfuge tube containing an aqueous solution containing internal standard. In the bioanalytical laboratory the sample was extracted using trichloroacetic acid, centrifuged and the supernatant available for instrumental analysis.

**Analytical methodology:**

A Shimadzu Nexera LC-MSMS was used to measure gentamicin concentrations, monitored in positive mode (ESI+) electrospray using selective reaction monitoring (SRM) transitions of 464.1→322.2, 450.1 →322.2 and 478.1→322.2. The internal standard (IS) kanamycin-B was monitored in ESI+ using SRM transitions of 484.05→205.1. An analytical column SeQuant ZIC-HILIC Guard, 20 x 2.1 mm was used for chromatographic separation with a total run-time of 3 minutes. Our batch analysis of samples included calibration standards, across a clinically-relevant concentration range and quality control samples, as well as drug-free matrix-matched samples.

**Reporting of results:**

Time to report gentamicin concentration data was recorded via electronic notification (using a short message service) from (1) the research bioanalytical laboratory and (2) the neonatal intensive care unit to a research study team member.
